# ALTARN: A Tabular Residual Neural Network for Alzheimer’s Disease Classification and Prediction

**DOI:** 10.1101/2025.10.21.25338451

**Authors:** Keshav Balakrishna, Alessandro Hammond, Abdeslem El Idrissi

**Affiliations:** University of Texas at Dallas; Department of Medicine, Harvard University; College of Staten Island, City University of New York

**Keywords:** Alzheimer’s Disease (AD), Attention Mechanism, Tabular Data, Sigmoid Weighting, Residual Neural Networks

## Abstract

Early and accurate prediction of Alzheimer’s disease (AD) from accessible clinical data remains a significant challenge in healthcare. This study proposes ALTARN, a tabular attention residual neural network architecture for robust classification of AD through heterogeneous patient data from a publicly available dataset of 2149 subjects, with medical, demographic, and lifestyle variables organized as structured tabular data. With sigmoid attention mechanisms for the dynamic reweighing of input variables for each patient, deep residual connections capturing complex, and non-linear relationships in tabular features, we propose ALTARN-an early Alzheimer prediction tool. ALTARN achieved an average cross-validated training accuracy of roughly 92.73%, alongside robust validation metrics, including an average accuracy of 85.06%, average precision of 80.73%, mean recall of 76.18%, and a mean validation F1-score of 78.32% when evaluated through five-fold stratified validation. When tested against other approaches, ALTARN meets and also exceeds the performance of supervised deep learning models for non-imaging AD classification. This further illustrates that deep neural network based approaches with tabular attention offer a promising direction for interpretable diagnosis of AD via non-imaging medical data.

## I. Introduction

Alzheimer’s disease (AD) is a progressive neuro-degenerative disorder that is marked by mild to severe losses in cognition and impairments in various domains of daily functioning. Such neurological impairment accounts for at least two-thirds of dementia cases in individuals aged 65 and older, and affects 43.8 million individuals as of 2016 [1]. Dementia is often a broader term used to describe a significant decline in cognitive ability that interferes with an individual’s activities of daily living (ADLs). Artificial intelligence (AI) and machine learning (ML) have been shown to be highly effective in their ability to enhance detection of AD in diverse arrays of patients. AI and ML has the ability to leverage subtle patterns and interactions to allow for earlier detection, improved prognosis, and more personalized treatment approaches to patient care. In particular, deep learning (DL) approaches, such as three-dimensional, DenseNet-169 and ResNet-50 convolutional neural network architectures, have previously demonstrated remarkable potential to enhance binary classification and detection of AD. Previous literature has shown a focus on the construction of reference models based on volume and thickness of established brain regions known to be responsible for disease progression [2][3]. Machine learning and deep learning methods for enhanced early classification of AD have demonstrated exceptional training and validation accuracy in the context of computer vision, wherein information must be regularly extracted from high dimensional data [4]. However, a number of obstacles are have been associated with a plateau of progress towards this goal via traditional supervised learning algorithms. Specifically, the classification of high dimensional data struggles due to homogeneity in existing small-scale data despite the heterogeneity of data in actual clinical applications and the overly supervised nature of pre-existing analysis methodology has hindered the development and promotion of a more novel, unsupervised analysis method [5].

Consequently, there is a increased necessity for more efficient and accurate classification methods for Alzheimer’s disease that can be integrated within mainstream clinical flows. Recent advents in deep learning have demonstrated the potential of tabular attention frameworks to enhance predictive accuracy and interpretability of existing supervised ML and DL diagnostic models. These frameworks improve model transparency and augment more nuanced features in tabular clinical data, which enables the prioritization or ranking classification of disease-sensitive variables with high precision. They have achieved remarkable accuracy gains over existing state-of-the-art DL neural network methods. As such this underscores the significance of attention-driven tools and their application to advance early AD prediction and clinical decision-making [6][7]. Currently, deep learning methods employ architectures such as VGG-16 with a fully connected layer, VGG-19 with specific frozen layers during training, and 3D ConvLSTM, among many others. When evaluated with cross-dataset validation, these models demonstrated high training accuracy but struggled to adapt to samples outside its distribution, which could potentially be explained by overfitting and reduced generalizability. An salient area to consider is the integration of existing DL analysis tools in clinical workflows: in reallife scenarios, variations in patient demographics and imaging protocols are inevitable. Hence, more advanced frameworks, e.g. a tabular attention mechanism, equipped with heightened sensitivity to heterogeneous data are necessary in order for AD predictive systems to be integrated into regular medical practice [8].

This paper proposes ALTARN, a novel tabular-attention residual neural network (RNN), trained on a multi-class dataset [12] such as medical, demographic, and lifestyle variables coupled with residual connections capturing complex relationships in tabular clinical features. When evaluated five-fold stratified validation, ALTARN delivers competitive performance, with an average validation and training accuracy of 85.1% and 92.73%, respectively. The proposed model achieved an average precision of roughly 80.73% and mean recall and F1-scores of 76.18% and 78.32%, respectively. Such results indicate a balanced performance across all stages of dementia and low false positive and false negative rates. ALTARN outperformed other models when compared with previous approaches in Huang (2023)[6]. As a consequence, the results demonstrate that a tabular-attention mechanism combined with a deep RNN can effectively learn discrete patterns that traditional and supervised neuroimaging-based approaches may misinterpret. Therefore, in conjunction with the sigmoid weighting attention mechanism, the results of this study underscore that a tabular-based RNN, which could potentially be a viable framework for the enhanced classification and prediction of AD via non-imaging medical data analysis.

## II. Literature Review

### A. Attention Mechanisms and Residual Networks in Medical Data Classification

Attention frameworks have emerged as powerful tools in medical data classification, and they are commonly used to sync to improve the accuracy and interpretability of previous deep learning analysis models as the frequency of overfitting towards training data is reduced. While significant neuroimaging approaches to AD detection exists, e.g. Magnetic Resonance Imaging (MRI) and Positron Emission Tomography (PET) scans, tabular data found in AD datasets often feature important biomarkers for diagnosis of the disease [2][6]. Huang (2023) proposed an attention mechanism to enhance and attribute salient features for the prediction of AD from deep learning models trained using tabular clinical data from 882 MR image scans located in the Alzheimer’s Disease Neuroimaging Initiative (ADNI) dataset, while implementing benchmarks featuring several supervised learning models. The model reached a validation accuracy of over 83.8%, a significant increase relative to classic neuroimaging-based models for AD classification and detection. Therefore, this suggests the flexibility of tabular-based attention mechanisms in the interpretability of patient clinical features [6].

Residual networks (ResNets) have also shown tremendous promise in the field of deep learning for medical data classification as they address the sub-optimal feature extraction and vanishing gradients in supervised deep learning models.

In particular, ResNets enable neural networks to learn identity mappings and allow for more advanced architectures to be trained that can capture a variety of scope representations-all of which are critical for nuanced medical data found in several Alzheimer’s disease clinical feature datasets. In AD classification, the integration of residual connections has been shown to improve diagnostic accuracy in Alzheimer’s disease classification. Zhang et al. (2022) introduced a 3D Residual Attention Deep Neural Network (ResAttNet) using structural MRI which conjoins residual learning and self-attention modules, achieving comparable and even superior accuracy, precision, and generalizability relative to conventional approaches [9]. In general, a residual self-attention framework improves the ability of a deep neural network to recognize salient brain regions™such as the cortex and hippocampus™and effectively automate the classification of AD in its various stages, underscoring the efficacy of ResNet architectures in medical classification tasks and its potential for mainstream applications [7].

### B. Deep Learning Approaches for Alzheimer’s Disease Prediction from Tabular Clinical Data

Deep learning approaches for the prediction of Alzheimer’s disease from tabular clinical data have gained considerable attention due to their versatile architecture: their ability to capture complex, non-linear relationships among diverse clinical features in non-homogeneous patient datasets. Newly emerging research highlights the efficacy of advanced deep learning models™equipped with fully-connected neural networks and attention-based mechanisms™in leveraging demographic, cognitive, biological, and neuroimaging-derived variations of tabular clinical data used for early detection of AD.

One such direction involves the integration of tabular clinical data with imaging and other crucial biomarker information using multi-modal deep learning to improve diagnostic accuracy of existing models. Qiu et al. (2022) proposed a multi-modal deep learning model collected clinical variables from neuroimaging data. Shapley Additive Planations proposed to be used for predictions with anatomical markers such as hippocampal and temporal lobe atrophy [10]. Recurrent neural networks have been show to successfully identify rapid AD progression with longitudinal tabular patient data (i.e brain volumes, neurocognitive scores, and more specific biochemical markers such as plasma amyloid-beta levels). Ma et al. (2024) found that these models achieved robust validation with AUROC from 0.75 to 0.83. It can be extrapolated the potential application of deep learning models to identify patients with a faster-than-average AD progression [11]. Data augmentation can be applied separately to tabular clinical features can enhance the robustness of existing classification models, while model depth peaks at an intermediate level with accuracy to balance overfitting and underfitting. Overall, multi-modal frameworks that combine clinical tabular data with biomarker information, recurrent neural networks with attention-focused mechanisms for longitudinal progression, and rigorous modeling choices advance reliable AD prediction tools for clinical deployment.

## III. Methodology

In this study, a publicly available dataset comprised of 2149 subjects is utilized in the training of ALTARN [12], which contains tabular biomedical records, where each row represents a unique individual. Specifically, each record contains variables such as demographics, lifestyle factors, clinical and family history, neuro-cognitive assessment scores, vital signs and laboratory measures, and finally an outcome label indicating the presence (1) or absence (0) of AD in each instance determined through clinical diagnosis. Additionally, as the proportions of AD-negative and AD-positive instances may not be balanced, the five-fold stratified validation framework in this paper is utilized to address the issue. Mathematically, for *m* patient features and *n* individuals:

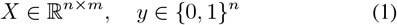

where *y* and *X* are the binary target and feature data, respectively. Then, in order to identify the dataset column that serves as the binary label and that is represent diagnostic outcomes, the proposed model searches through columns with only two or three unique values. In particular, all unique values in the target candidate column are mapped and checked to the binary set {0, 1} where string values such as {Yes, true} correspond to the presence of AD, while values such as {No, false} correspond to an absence of AD. If a value is missing, the label is imputed as 0 to maintain a conservative approach in biomedical settings where:

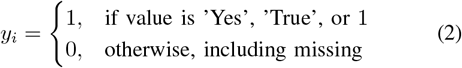

This process thus ensures a numerically encoded and clean outcome vector compatible with ML frameworks, and simultaneously prevents inconsistent downstream interpretations. As most neural networks and ML frameworks require numeric input data, and in order to be lossless for categorical information, all non-numeric and non-target features (e.g. ethnicity and education) are label encoded such that every unique category is represented as an integer, mathematically represented as:

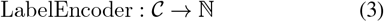

where 𝒞 represents the set of unique categorical values for a given feature. If a feature value is missing in any sample, all unknown values are assigned to a special label as part of encoding for categorical features, whereas missing entries are replaced with the mean of the feature (computed from the available data), mathematically denoted as:

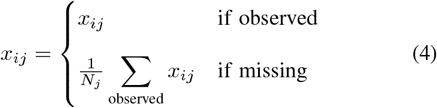

where *x*_*ij*_ denotes the value for feature *j* and sample *i*, which preserves as much information as possible, while it also prevents dropping samples and allows for sensible estimates for absent values. Next, to ensure equitable feature contribution and efficient convergence of the learning algorithm, all numerical features are standardized using a *z*-score normalization. The procedure to scale centers the data around zero and normalizes the variance to one. The transformation is expressed as:

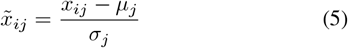

where *µ*_*j*_ is the mean and *σ*_*j*_ is the sample standard deviation for feature *j*. Standardization allows for stable training, especially for deep architectures with gradient based optimization. After it is preprocessed, each sample is represented by a fully numeric standardized feature vector that serves as an input to the ALTARN model. The vector is mapped with a dense layer to a latent representation of dimension 128. Batch normalization and rectified linear activation is used to ensure nonlinearity and stable gradients:

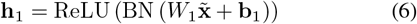

On this latent encoding, a feature wise mechanism is employed through the use of a learnable weight. The learnable weight is computed for each input variable via a sigmoid transformation so the network can dynamically focus on clinically salient features for each prediction:

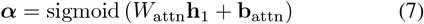

The input is then formed through element wise multiplication,

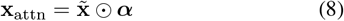

such that aforementioned computations exert greater influence due to variables being more informative. The model also employs a residual learning block to allow both expressive transformation and preservation of salient low-level patterns. The attended vector gets processed through two parallel paths: a dense layer (dimension 64) followed by ReLU activation and dropout regularization. The sum of the resulting outputs creates the next latent vector:

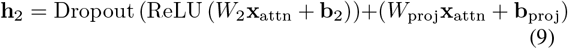

Such a design allows for stability in deep learning while guarding against information degradation when signals propagate through layers. Further processing involves a dense layer (of size 32) with additional dropout and non-linear (ReLU) activation applied to **h**_2_ to produce the latent vector **h**_3_:

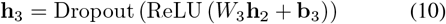

A final neuron with sigmoid activation then produces the probability 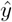 of Alzheimer’s diagnosis, where **h**_3_ represents the output applying a dense layer (width 32), followed by a ReLU activation and dropout to **h**_2_:

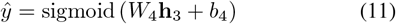

A probability greater than 0.5 is interpreted as a positive diagnosis for Alzheimer’s disease, while lower values indicate a negative result. To train the model, binary cross-entropy is used as the loss function to guide optimization.

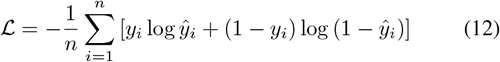

Afterwards, the Adam Optimizer is utilized due to its adaptive learning rate and robust convergence properties in deep learning tasks. The model is trained with a batch size of 32 and for a maximum of 200 epochs. Early stopping is employed with a patience of 10 epochs to limit overfitting. Once the validation loss ceases to improve, the model training will abruptly end and automatically restore the weights from the epoch with the best validation performance. To rigorously evaluate generalizability, the study deploys stratified five-fold cross-validation. This allows the proportion of AD-positive and AD-negative cases to be consistent in each partition with the training and validation. For each fold, accuracy, F1 scores, precision, and recall were computed and then averaged across the five folds.

## IV. Results and Analysis

Evaluated and trained through five-fold stratified validation, ALTARN achieved the training and validation results across all folds, as shown by the table below:

As illustrated by the results in Fig. 2, ALTARN achieves an average training accuracy of 92.73% and mean validation accuracy of roughly 85.06%. Furthermore, with validation F1-scores averaging to roughly 78.32%, the model achieves a balanced performance across all dementia stages, despite class imbalance, and the consistency between all folds illustrates robustness and generalizability. Finally, validation precision and recall metrics, of 80.73% and 76.18%, respectively, indicate both low false positive and false negative rates, further illustrated by the results of Fig. 4. Based on the results of the confusion matrix portrayed in Fig. 3, the model correctly predicted 246 instances in Class 0 that were truly in Class 0 (true negatives) and 108 samples in Class 1 that were truly in Class 1 (true positives), while it predicted incorrectly 31 samples as part of Class 1, when in reality in Class 0 (false positives), and 44 samples in Class 0 when they were truly in Class 1 (false negatives). Due to the higher number of predicted instances towards the main diagonal (top left and bottom right), alongside lower amounts for the off-diagonal values (bottom left and top right), the model demonstrates robust performance in accuracy. To contextualize the results of ALTARN, as depicted in Fig. 4, the model is benchmarked to similar approaches in AD classification using tabular medical data:

**Fig. 1.**
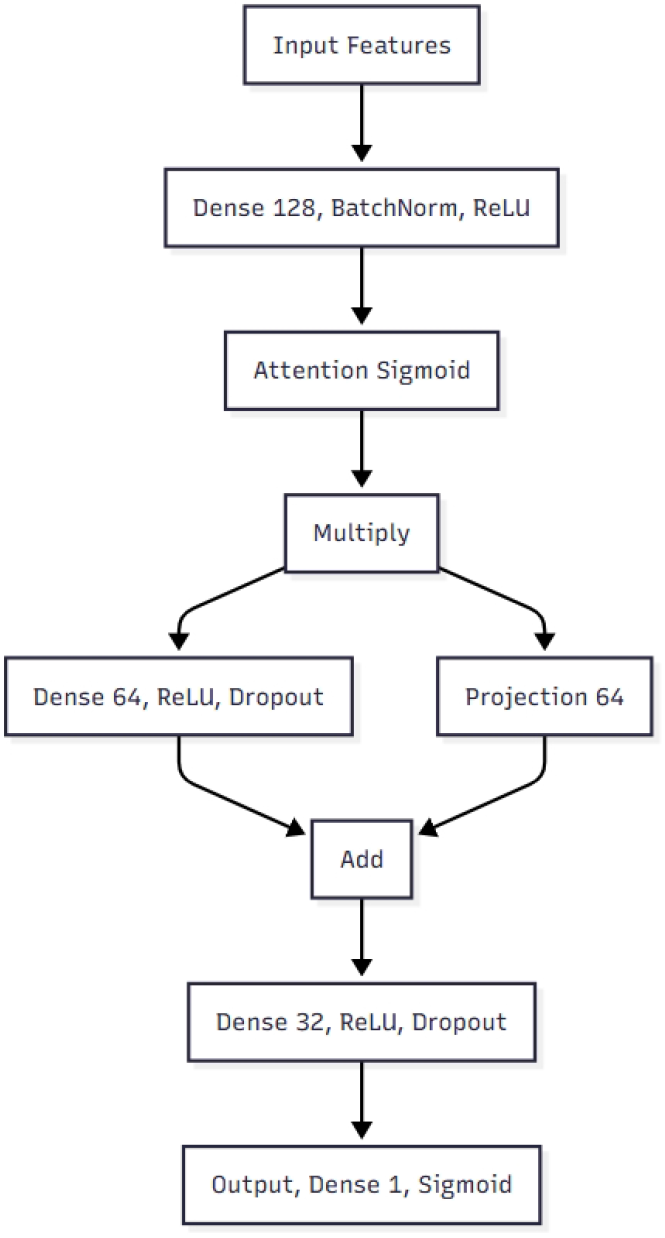
ALTARN Architecture Diagram

**Fig. 2.**
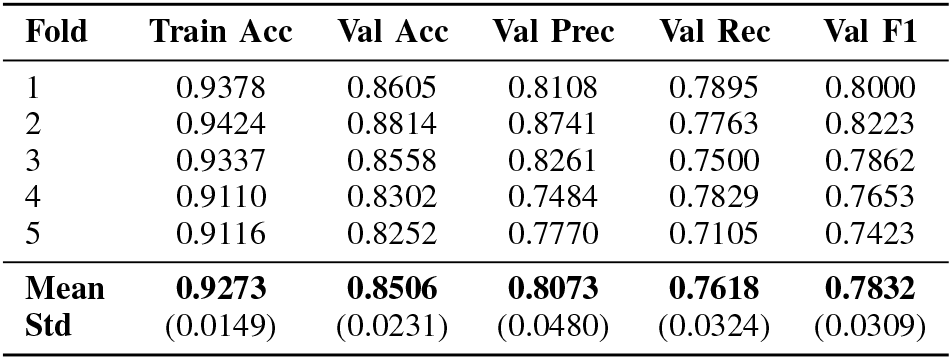
5-fold stratified cross-validation results of ALTARN on Alzheimer’s tabular data (2149 subjects). Metrics reported as mean *±* standard deviation.

**Fig. 3.**
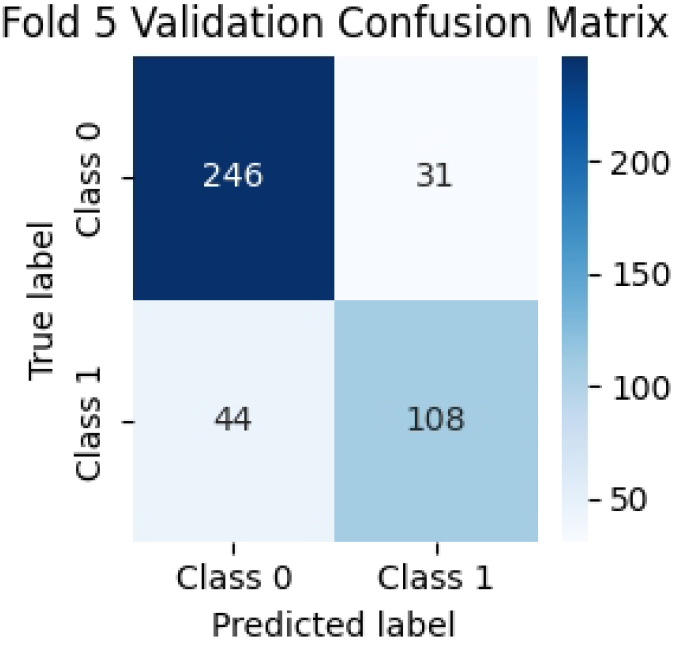
Fold 5 Cross-Validation Confusion Matrix

**Fig. 4.**
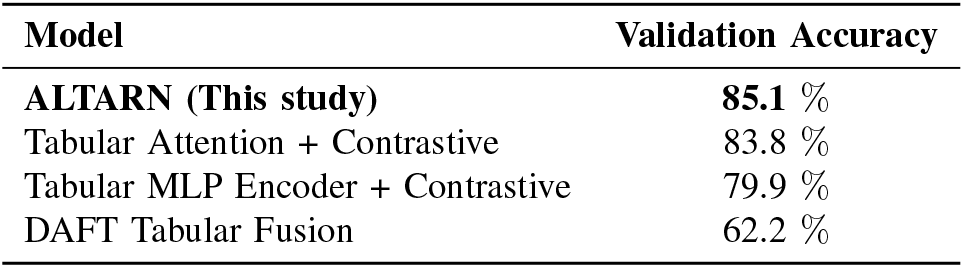
Validation accuracy comparison of ALTARN and recent deep tabular architectures [6]

As illustrated in Fig. 4, ALTARN has comparable performance in comparison to other similar approaches, such as Tabular Attention + Contrastive and DAFT Tabular Fusion.

Overall, the combination of sigmoid attention and residual connections ensures that the network focuses on the most salient features relevant to multi-stage AD detection in clinical MRI scans. Furthermore, robust validation metrics illustrate that ALTARN could be viable for clinical decision-support pipelines and offer reliable differentiation among the different severity stages of AD, with the consistent fold performance underscore a robust potential in varying clinical data splits.

## V. Conclusion and Discussion

This study proposes ALTARN, a Tabular Attention Residual Neural Network framework, for the prediction of AD through structured clinical tabular data, and integrates the Sigmoid attention mechanism directly on input features alongside residual connections to enhance both learning capacity and spatial recognition in tabular data structures. Evaluated through five-fold stratified validation on a publicly available clinical AD dataset, ALTARN achieved a robust performance with an average cross-validated training accuracy of roughly 92.73%, alongside validation metrics, including an average accuracy of 85.06%, average precision of 80.73%, mean recall of 76.18%, and a mean validation F1-score of 78.32%. As a consequence, the results suggest that ALTARN effectively learns discriminative patterns in clinical features which correlate with disease status and maintains robust generalization to unseen instances. In addition, the low standard deviation across all folds illustrate a reliable, consistent performance, which demonstrate model stability regardless of data split variation. Therefore, the findings emphasize the potential of the combination of residual learning and attention in neural network architectures tailored for tabular medical datasets, where traditional deep learning approaches often struggle in comparison to sequential or imaging tasks. Most notably, ALTARN achieved competitive performance against several ML-based approaches on similar AD classification tasks, thus supporting its use in clinical decision support systems that aim to stratify patient risk through multi-modal clinical features. Despite these promising results, there are several limitations: first, only one limited dataset with a potentially restricted demographic or clinical diversity is utilized, which could reduce the generalizability of the model to broader populations and heterogeneous clinical environments. While ALTARN integrates an attention mechanism to produce salient input features, no interpretability analyses or explainable AI (XAI) techniques (i.e feature importance ranking or SHAP values) were used, which can affect regulatory approval and model rationale in decision support contexts. Error analysis was limited, and the model’s misclassifications were not systematically examined across different clinical subgroups and stages of AD. Future work should address these limitations by with rigorous XAI methods to elucidate model decision-making, and perform detailed error analyses to identify systematic biases.

## Data Availability

All data produced in the present work are contained in the manuscript.

## VI Acknowledgements

The authors thank Abhiram Yaramsetti for his valuable support and contributions.

